# Oromucosal administration of oxytocin and its receptor antagonist atosiban respectively increase and decrease preferential attention to dynamic social stimuli in men

**DOI:** 10.64898/2026.09.08.26362314

**Authors:** Yan Zhang, Xinling Wang, Yige Wang, Huan Liu, Keith M. Kendrick

## Abstract

**Background:** Reduced concentrations of the neuropeptide oxytocin can occur in disorders with social dysfunction, notably autism. However, the role of peripheral endogenous oxytocin concentrations and increased ones following exogenous administration in modulating key social behaviors in humans is still unclear. Here we investigated effects of targeting peripheral oxytocin receptors through oromucosal administration of oxytocin and an oxytocin receptor antagonist (atosiban) on social attention.

**Methods:** We carried out a pre-registered, randomized, double-blind, placebo-controlled experiment with 250 adult male subjects in five treatment groups (placebo + placebo, placebo + 24 IU OT, placebo + 48 IU OT, atosiban + placebo, and atosiban + 24 IU OT), each receiving two oromucosal (lingual) administrations separated by 15 minutes. Following treatment, subjects performed an autism-sensitive dynamic social versus geometric pattern eye-tracking paradigm. Pre- and post-treatment blood samples were collected for oxytocin measurement.

**Results:** Results showed that 48IU but not 24IU oxytocin significantly increased preferential viewing of social stimuli relative to placebo, and effects were associated with, and mediated by, increased oxytocin concentrations and negatively correlated with subjects’ autism quotient scores. On the other hand, atosiban reduced preference for social stimuli relative to placebo even after subsequent administration of 24IU oxytocin.

**Conclusions:** Our findings suggest that both exogenous and endogenous oxytocin can act via peripheral receptors to enhance interest in social relative to non-social stimuli, possibly influencing the brain via acting on G[q]-coupled receptors in the vagal system. This further supports the therapeutic potential of oxytocin for social dysfunction.

## Introduction

Oxytocin (OT) is a hypothalamic neuropeptide demonstrated to play a key role in regulating social attention, cognition, motivation and behavior across species (Meyer-Lindenberg et al., 2011; Kotani et al., 2017; Yao & Kendrick, 2025). In humans, clinical studies have frequently reported decreased endogenous concentrations of OT in psychiatric disorders with social dysfunction such as autism spectrum disorder (Moerkerke et al., 2021), depression (Xie et al., 2021) and schizophrenia (Ferreira & Osório, 2022), although with some age or sub-group dependent effects. However, it is unclear what the importance of basal endogenous concentrations is for social behavior and while numerous studies in both healthy and clinical populations have reported facilitation of various aspects of social behavior following exogenous administration by an intranasal route, findings have often been inconsistent and the mechanisms involved are not fully understood (Yao & Kendrick, 2025). For exogenous administration, studies have primarily adopted intranasal administration of OT based on claims that it can enter the brain via this route. However, several studies have shown that OT does not cross the blood-brain barrier in appreciable amounts (Kendrick et al., 1986; Mens et al., 1983) and while some studies have suggested it may enter the brain via the olfactory and trigeminal nerves (Lee et al., 2020), others have shown limited to no target coverage of central OT receptors at the doses typically used based on cerebrospinal concentrations in humans and non-human primates (Bowen, 2019). Other research in rodents, has suggested OT can cross the blood-brain-barrier after binding to the receptor for advanced glycation end-products (RAGE) (Yamamoto & Higashida, 2020) but so far there is no evidence for this in humans. Overall, recent studies have increasingly demonstrated that functional effects of OT following intranasal administration may primarily be mediated via it entering into the peripheral circulation and acting on [OT receptors](https://www.guidetopharmacology.org/login.jsp) in the heart and gastrointestinal systems to influence the brain via vagal projections to influence activity in regions of the social brain directly and/or via promoting OT release within the brain (Yao & Kendrick, 2025; Zhu et al., 2022) in support of Porges original polyvagal theory of the importance of interactions between the peripheral autonomic system and brain via the vagus for controlling socioemotional behavior (Porges, 2021). Thus, reducing the ability of intranasal OT to increase peripheral concentrations in humans prevents it influencing resting state neural changes (Yao et al., 2023). Furthermore, administering OT via oral routes (oromucosal: lingual or medicated lollipop), which is unlikely to result in it directly entering the brain in humans, increase plasma OT concentrations in 15 min (Xu et al., 2022) and can produce similar effects on social behavior as intranasal administration (Xu et al., 2024; Kou et al., 2021; Chen et al., 2022). These functional effects of oromucosally administered OT are also partially mediated by the increase in peripheral concentrations of the peptide (Kou et al., 2021; Chen et al., 2022). However, it is still to be fully demonstrated if exogenously administered OT is indeed producing its effects via acting on its peripheral receptors.

An important approach in terms of establishing potential functional effects of either basal endogenous OT concentrations or increased ones following exogenous administration is to investigate the impact of competitive OT receptor antagonists which can prevent OT from binding to its peripheral receptors. Atosiban is a competitive antagonist of OT receptors that exhibits strong and specific binding to the OT receptor, weaker binding to the V1a receptor (V1aR), and comparatively less specific binding to the V2 receptor (V2R) (Reversi et al., 2005; Ślusarz et al., 2004). While atosiban acts primarily as a competitive antagonist on OT receptor G(q) coupling, it is a partial agonist on G(i) coupling (Reversi et al., 2005). Peripherally administered atosiban is not thought to cross the blood brain barrier in appreciable amounts (Marshall, 2007) and thus any functional effects it has on social behavior are likely to be primarily as a result of preventing OT from acting on peripheral receptors. To date, intranasal atosiban has been reported to influence the perception of time in social interactions (Liu et al., 2018) and reduce the ability to discriminate steroid components of male and female odors relative to OT (Chen et al., 2021). However, intranasal administration of atosiban may have potentially influenced either central or peripheral OT receptors or both, similar to intranasal oxytocin. On the other hand, if atosiban is administered oromucosally, it is likely mainly to only act as a competitive antagonist on peripheral OT receptors.

One of the key functional effects of OT is facilitation of attention to salient social cues, using a number of different paradigms (Yao & Kendrick, 2025). A paradigm which has received increasing attention is also particularly sensitive for discriminating autistic from neurotypical individuals (Kou et al., 2019; Moore et al., 2018; Pierce et al., 2016). This paradigm uses eye-tracking to measure visual attention towards paired dynamic video stimuli featuring individuals dancing or geometric patterns, with neurotypical individuals exhibiting greater interest in the social stimuli whereas autistic individuals are more interested in the geometric ones. This is sometimes referred to as the ‘GeoPref’, preference for dynamic geometric stimuli, paradigm (Pierce et al., 2016). Acute, intranasal OT can increase the time spent looking at the social stimuli in non-autistic adults (Le et al., 2020) and chronic intranasal OT administration (6 weeks, every other day) can also do so in young autistic children (Le et al., 2022). However, the functional importance of OT acting on peripheral receptors, whether at basal concentrations or at increased concentrations after oromucosal administration, has yet to be evaluated in this paradigm.

In the current study, we therefore investigated the respective effects or oromucosal (lingual) administration of OT and its receptor antagonist atosiban on the performance of adult male subjects in the above dynamic visual attention paradigm. Five different treatment groups were included where two treatment phases were incorporated primarily to allow atosiban to be administered prior to OT. The two treatments were separated by 15 min and included placebo + placebo; placebo + 24IU OT; placebo + 48IU OT; atosiban + placebo and atosiban + 24IU OT with subjects performing the paradigm 30 min after the second treatment phase. Given previous evidence that OT-effects in this paradigm are influenced by autistic traits, correlations between behavioral performance and autistic traits were also analyzed. A 48IU dose was included both to provide a dose response analysis and also because 24IU OT given orally raises OT concentrations significantly less than the same dose given intranasally (Xu et al., 2022; Xu et al., 2024). Since atosiban is a competitive antagonist, and to avoid further increasing group numbers, we chose to administer it only prior to the lower dose of OT as we predicted it would be more likely to reduce or block OT effects by doing so. Blood samples were also taken before and after each treatment for measurement of OT.

We hypothesized that OT would dose-dependently increase the time subjects spent viewing dynamic social stimuli and that atosiban would have the opposite effect if acting primarily as a G(q) channel antagonist. We also hypothesized that pre-treatment with atosiban would reduce the effectiveness of 24IU OT, that effects of OT would negatively correlate with trait autism scores and that the magnitude of changes in plasma OT would positively correlate with time spent viewing social stimuli.

## Materials and methods

### Participants and experimental procedure

A total of 250 adult male participants were recruited for the randomized, double-blind, placebo (PLC)-controlled study with 50 participants assigned to five different treatment groups using a computer-generated randomization procedure. Individuals in each group received two treatments separated by 15 min (PLC + PLC; PLC + 24 IU OT; PLC + 48 IU OT; atosiban (150μg) + PLC; atosiban (150μg) + 24 IU OT; see Fig. 1 for experimental protocol). PLC, 24IU OT, 48IU OT, atosiban, atosiban + 24IU OT were used as the abbreviations for the respective groups. Based on an a priori power analysis using G*Power for a two-way mixed analysis of variance (ANOVA), the sample size was adequate to achieve a power >0.8 (medium effect size f = 0.25, α = 0.05). Each treatment was administered as 6, 0.1 ml sprays (alternating supralingual and sublingual) with each spray separated by 30 s and participants instructed not to swallow during the 30 s after each spray to help maximize vascular absorption. Doses of 24IU and 48IU OT (oxytocin acetate – synthetic, supplied by Sichuan Defeng, Pharmaceutical Company, Chengdu, China) were given diluted in 0.9% sodium chloride and glycerol with placebo doses only including the diluent. Atosiban (atosiban acetate – synthetic, Sigma-Aldrich, St. Louis, MO, USA) was also prepared as a sterile solution diluted using the placebo solution and a 150μg dose was chosen based on two previous studies which administered 60μg intranasally (Liu et al., 2018; Chen et al., 2021). We used a higher dose given that greater amounts of substances may enter the peripheral circulation with an intranasal relative to a lingual route (Xu et al., 2022; Xu et al., 2024). To avoid possible confounds by administering different numbers of sprays, concentrations were adjusted so that doses were all given in 6, 0.1ml sprays. To confirm the success of blinding, after the experiment all participants were asked to guess which of the three different potential treatments they had received (i.e. placebo / OT / atosiban) for each of the two sprays. Multiple analyses showed that subjects in all treatment groups were unable to accurately identify which treatments they had received (see Supplementary methods for details). All participants were instructed to abstain from caffeine and alcohol for 24 hours and fast for two hours before the experiment. Exclusion criteria included self-reported: (i) History of or current neurological/psychiatric disorders; (ii) Use of psychotropic medications (including nicotine). 23 participants were excluded from analysis due to: failure to complete the experiment (*n* = 2), incomplete data (*n* = 8); insufficient attention (< 70% of total time spent viewing the stimuli; *n* = 13). Thus, the final analyses included data from 227 male participants (mean age = 21.14 years, *SD* = 2.21).

**Fig 1.**
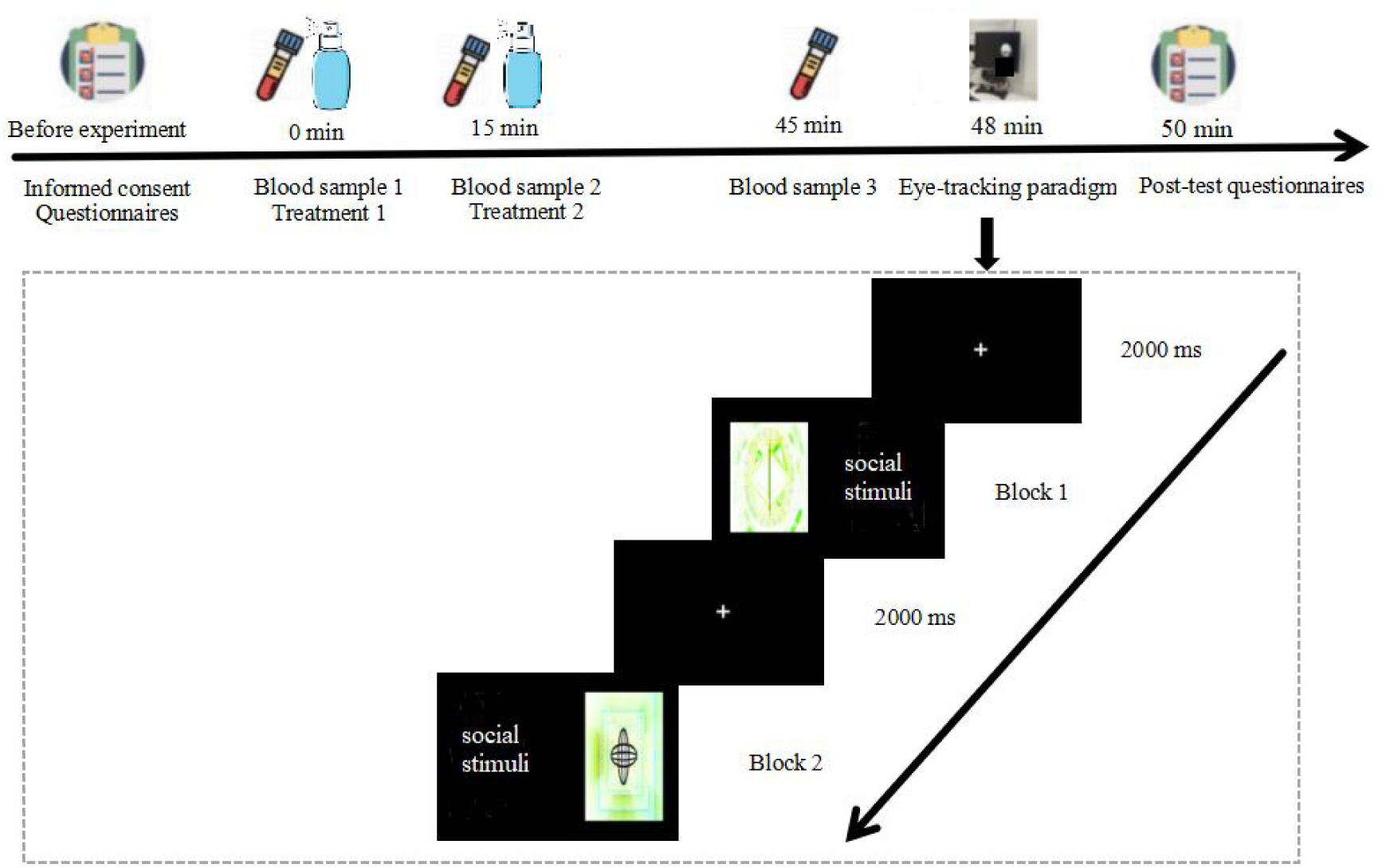
Experimental protocol. Top shows the time-line of the experiment for completion of questionnaires, lingual treatments, blood samples and performance of the eye-tracking paradigm. The dynamic eye-tracking paradigm is illustrated below, given as two video blocks.

All participants completed Chinese versions of questionnaires for state-trait anxiety inventory (STAI) (Shek, 1993), Leibowitz social anxiety scale (LSAS) (Kubota et al., 2016), Beck depression inventory-Ⅱ (BDI-Ⅱ) (Yang & Stewart, 2020) and Autism Spectrum Quotient (AQ) (Lau et al., 2013) before the experiment. The state anxiety component of the STAI was administered both before and after (after completing the behavioral paradigm) treatments to assess potential treatment/paradigm effects on anxiety. Blood (via an indwelling median cubital vein catheter) samples were collected immediately before and 15 min after the first treatment and again 30 min after the second treatment, centrifuged and the plasma stored at -70C until assayed for OT. The 30 min time point post-treatment for blood collection and start of the experimental paradigm corresponds closely to our previous estimate of a Tmax of 34.5 min and subsequent duration of significant increase for another 30-45 minutes after 24IU oromucosal OT, with bioavailability calculated to be 4.45% (Kou et al., 2021; Xu D et al., 2024). In the intervals between treatments and the start of the behavioral paradigm participants remained in the lab and were asked to sit comfortably and relax (no mobile phone use or conversation with experimenters). The experimental protocol is shown in Fig 1.

The experiment was approved by the ethics committee of the University of Electronic Science and Technology of China and pre-registered as a clinical trial (NCT07093060). All participants provided informed written consent after receiving details of the experimental procedures.

### Behavioral task and equipment

The dynamic social versus geometric stimuli paradigm (DSG) was used as in our previous studies (Kou et al., 2019; Le et al., 2020). In brief, the presentation consisted of two blocks lasting a total of around 72 seconds. Each block presented social stimuli (each showing 18 different individual adults dancing around) and 18 constantly evolving dynamic geometric patterns. The location of the social and geometric videos was randomly presented either on the left or right side of the screen. This free-viewing task involved no instructions from experimenters or individuals in the videos, allowing for an unguided assessment of subjects’ spontaneous attention preference. Eye movement data were recorded using an EyeLink 1000 Plus system (SR Research, Ottawa, ON, Canada) in monocular mode with a 2,000 Hz sampling rate and a 1,024 × 768 screen resolution. The standard distance from participant’s eyes to the screen was fixed at 57 cm via a chin rest, and a nine-point calibration was performed before the paradigm to ensure high-quality eye-tracking data. Raw eye movement data were exported and pre-processed using EyeLink Data Viewer 3.1.

### Oxytocin assay

OT concentrations in plasma samples (1.2 ml) were measured in duplicate as in a number of our previous studies (Yao et al., 2023; Xu et al., 2022; Kou et al., 2021; Chen et al., 2022) using an ELISA (Enzo Life Science, Farmingdale, NY, USA, Cat# ADI-901-153A, RRID: AB_2815012) following 4-fold concentration and extraction steps. Sensitivity was 2 pg/ml with inter- and intra-coefficients of variation of < 10%. A total of 213 out of the 227 participants had OT concentrations successfully measured in all three samples and were included in the final analysis.

### Statistical analysis

Mixed ANOVA was employed to examine group differences in fixation time duration and number of fixations (fixation counts) for the two areas of interest (AOIs – i.e the dynamic social videos and the dynamic geometric pattern ones). For the two-level AOI factor, no sphericity correction was needed. The assumption of homogeneity of variance was assessed using Levene’s test. The same method was used to explore group differences in plasma OT concentrations over time. Because Levene’s test indicated no significant violation of the homogeneity of variance assumption for all dependent variables (all *ps* > 0.05), post-hoc tests were conducted using Tukey’s honestly significant difference (HSD) test with bootstrap 2,000 resamples to improve the robustness of estimation. A significance level of *p* < 0.05 was set as the criterion for statistical significance. Associations between the time spent viewing the social stimuli and peripheral concentrations of OT, and AQ scores were assessed using Pearson correlation with bootstrap 2,000 resamples to enhance result reliability. The mediation analysis was performed using the Process V4.1 plugin with bootstrap 5,000 resamples. All the statistical analyses were performed using SPSS 27.0.

### Nomenclature of Targets and Ligands

Key protein targets and ligands in this article are hyperlinked to corresponding entries in http://www.guidetopharmacology.org, and are permanently archived in the Concise Guide to PHARMACOLOGY 2025/26 (Alexander et al., 2025).

## Results

### Demographics and questionnaires

ANOVA analyses showed no significant differences between the 5 treatment groups in terms of age and questionnaire scores (see Table 1). Two-way mixed ANOVA analysis showed that state anxiety scores were significantly reduced post-relative to pre-treatment (*F* = 37.230, *p* < 0.001, ƞ_p_*^2^ =* 0.144) but there was no main effect of group (*F*= 1.690, *p* = 0.153, ƞ *^2^ =* 0.030) or time x group interaction (*F* = 1.096, *p* = 0.360, ƞ_p_*^2^ =* 0.019).

**Table 1.**
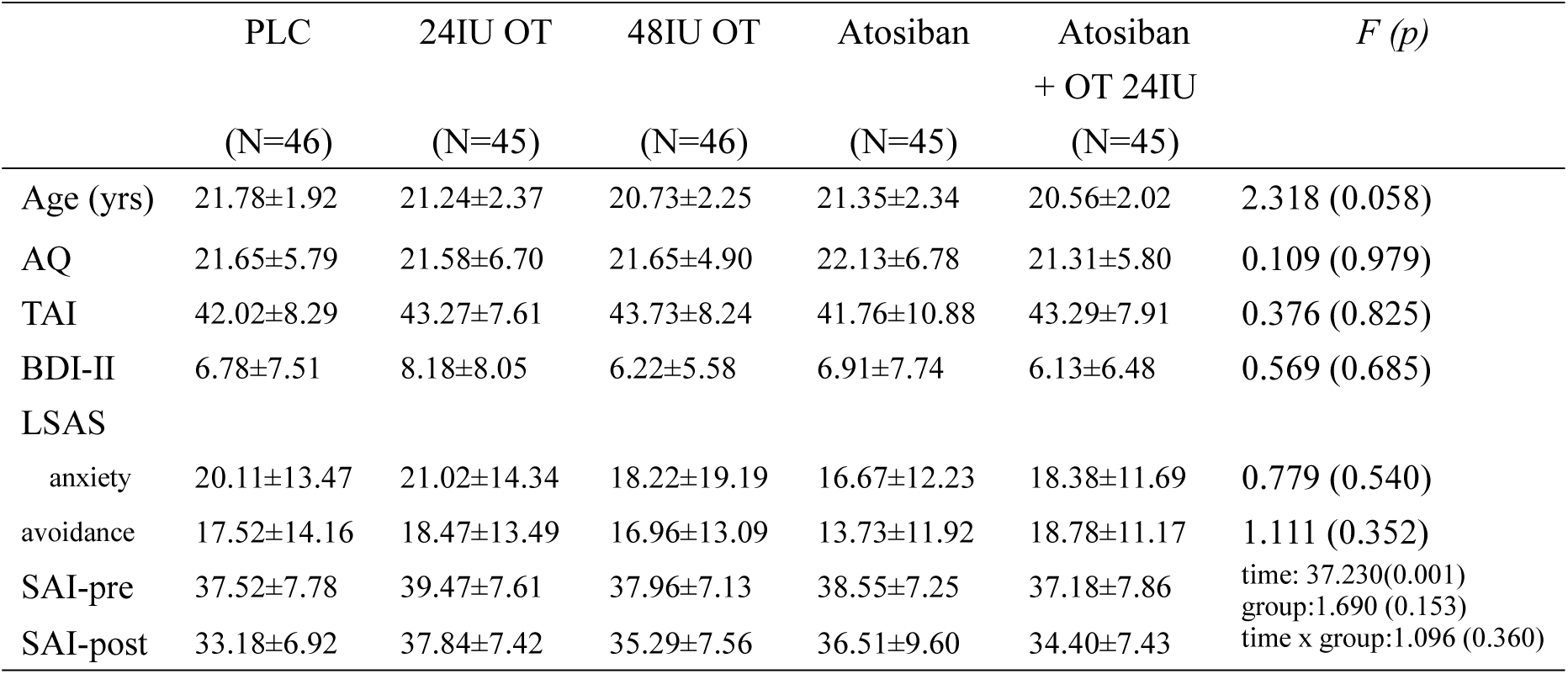
Participant demographics and questionnaire scores.

### Comparison of fixation durations and counts across the different treatment groups

Two-way mixed ANOVA analysis of fixation durations revealed no significant main effect of group (*F*(4,222) = 1.795, *p* = 0.131, ƞ_p_*^2^ =* 0.031), with all groups spending > 95% of the time viewing the videos, but a significant main effect of stimuli (*F*(1,222) = 70.920, *p* < 0.001, ƞ_p_*^2^ =* 0.242) and a group × stimuli interaction (*F*(4,222) = 7.000, *p* < 0.001, ƞ_p_*^2^ =* 0.112). The main effect of stimuli reflected greater fixation time on social relative to geometric stimuli across the groups (see Fig. 2). For the interaction, post-hoc tests corrected for multiple comparisons (Tukey) with 2,000 bootstrap samples revealed that fixation time was significantly greater for social stimuli in the 48IU OT group (*p* = 0.034, Cohen’s *d* = 0.422), but not the 24IU OT group (*p* = 0.745, Cohen’s *d* = 0.061) relative to PLC. On the other hand, the fixation time for social stimuli was significantly reduced in the atosiban group compared to PLC (*p* = 0.013, Cohen’s *d* = 0.483), 24IU OT (*p* = 0.030, Cohen’s *d* = 0.422) and 48IU OT (*p* < 0.001, Cohen’s *d* = 0.905) groups. The atosiban+24IU OT group had reduced fixation time on social stimuli compared to the PLC (*p* = 0.002, Cohen’s *d* = 0.545), 24IU OT (*p* = 0.012, Cohen’s *d* = 0.483) and 48IU (*p* < 0.001, Cohen’s *d* = 0.966) groups but not the atosiban alone group (*p* = 0.654, Cohen’s *d* = 0.061) (see Fig. 2). For fixation time on geometric stimuli the post-hoc analysis showed that this was reduced by 48IU OT (*p* = 0.014, Cohen’s *d* = 0.446) but not 24IU OT (*p* = 0.939, Cohen’s *d* = 0.022) compared to PLC (see Fig. 2). The fixation time for geometric stimuli showed a trend toward increase in the atosiban compared to PLC (*p* = 0.058, Cohen’s *d* = 0.397) and 24IU OT (*p* = 0.071, Cohen’s *d* = 0.375) groups and achieved significance compared to the 48IU OT group (*p* < 0.001, Cohen’s *d* = 0.843). The atosiban + 24IU OT group showed an increase in viewing time of geometric stimuli compared to the 48IU OT group (*p* < 0.001, Cohen’s *d* = 0.806) but not to other groups (*p* = 0.082, Cohen’s *d* = 0.361 vs PLC; *p* = 0.093, Cohen’s *d* = 0.338 vs 24IU OT; *p* = 0.869, Cohen’s *d* = 0.037 vs atosiban).

**Fig 2.**
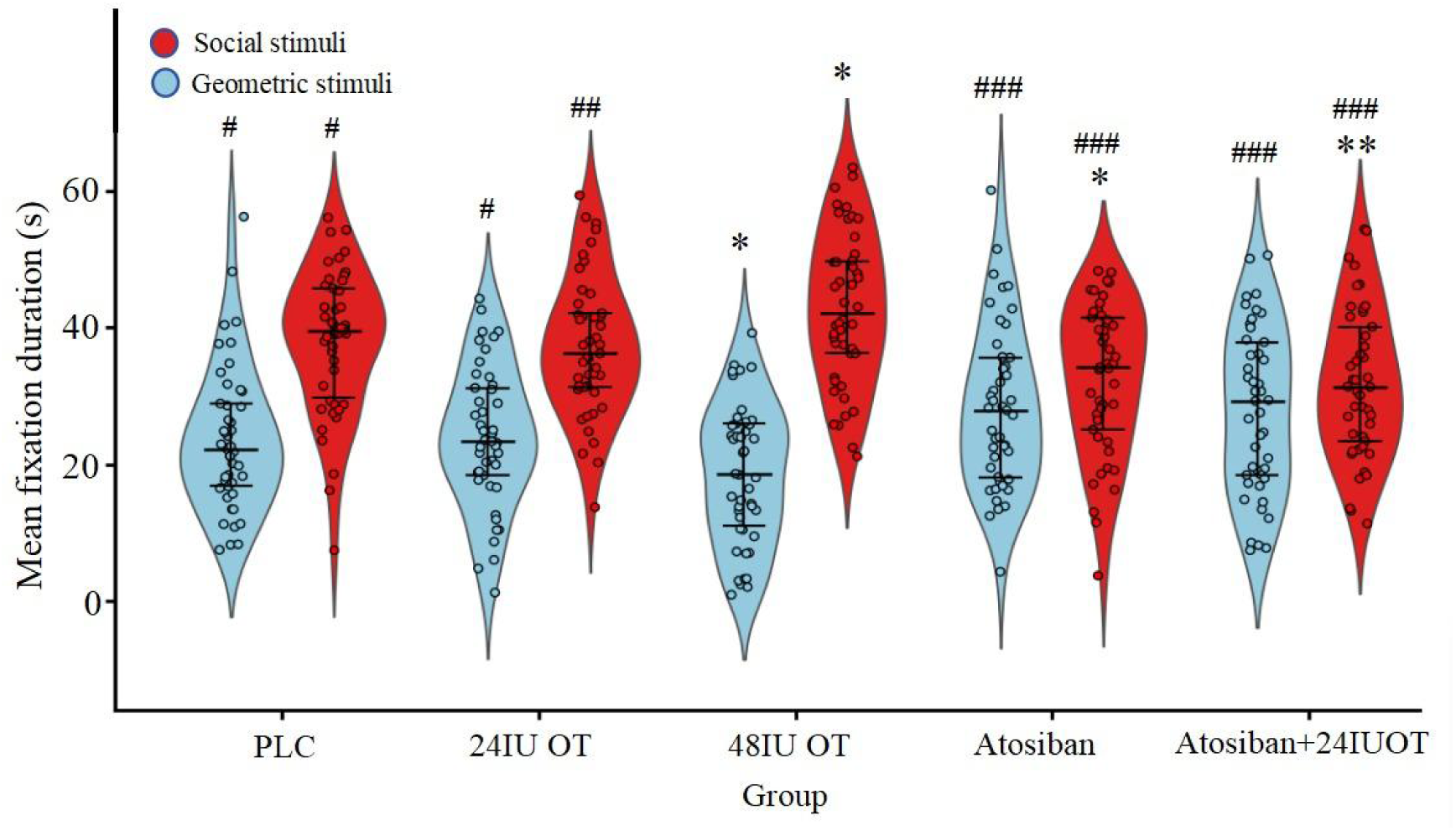
Treatment effects on time spent viewing social and geometric stimuli in the eye-tracking paradigm. Violin plots show individual and group data for time spent viewing either the dynamic social (dancing individuals) or non-social (geometric patterns) stimuli in the visual attention paradigm in the five different treatment groups. Lines within the violins also indicate interquartile ranges. \**p* < 0.05, ** *p* < 0.01, *** *p* < 0.001 compared to the same stimuli in the PLC group. #*p* < 0.05, ## *p* < 0.01, ### *p* < 0.001compared to the same stimuli in the 48IU group.

The same analysis of fixation counts revealed a main effect of stimuli (*F*(1,222) = 165.434, *p* < 0.001, ƞ_p_*^2^ =* 0.427) and also a stimuli x group interaction (*F*(4,222) = 5.038, *p* < 0.001, ƞ_p_*^2^ =* 0.083-see Supplementary Information Table S1) indicating that the greater overall fixation duration on social stimuli observed for OT 48IU was mainly contributed to by an increased number of fixations (*p* = 0.068, Cohen’s *d* = 0.349 vs PLC; *p* = 0.030, Cohen’s *d* = 0.441 vs 24IU OT; *p* < 0.001, Cohen’s *d* = 0.782 vs atosiban; *p* < 0.001, Cohen’s *d* = 0.804 vs atosiban + 24IU OT).

### Plasma OT concentrations across treatment groups and associations with social attention

To investigate absolute changes in OT concentrations across groups, we used the differences between T1 (basal concentrations) and T2 (times +15 min), and T3 (+ 45 min) as the dependent variables since this controls for the influence of individual differences in baseline concentrations. Two-way mixed ANOVA on changes in OT concentrations revealed that there were significant main effects of group (*F*(4,208) = 8.012, *p* < 0.001, ƞ_p_*^2^ =* 0.133) and time (*F*(1,208) = 45.214, *p* < 0.001, ƞ_p_*^2^ =* 0.179) and a significant interaction effect between group and time (*F*(4,208) = 13.935, *p* < 0.001, ƞ_p_*^2^ =* 0.211). Post-hoc tests showed there were no significant group differences in OT concentration changes at T2 (see Fig. 3a), but there were at T3 (T2: *F*(4,208) =0.755, *p* = 0.556, ƞ_p_*^2^ =* 0.014; T3: *F*(4,208) = 13.157, *p* < 0.001, ƞ_p_*^2^ =* 0.202). Post-hoc tests corrected for multiple comparisons (Tukey) with 2,000 bootstrap samples at T3 showed that OT concentrations in the 48IU OT group increased significantly relative to the PLC, 24IU OT, atosiban alone and atosiban + 24IU OT groups (all *ps* < 0.05). The OT changes were significantly higher in the 48IU OT group (*mean ± SD*, +6.66 ± 6.38 pg/ml) than 24IU OT (+3.51 ± 4.74 pg/ml, *p* = 0.006, *d* = 0.544), PLC (+0.06 ± 5.17 pg/ml, *p* < 0.001, *d* =1.139), atosiban + 24IU OT (+3.41 ± 5.66 pg/ml, *p* = 0.016, *d* = 0.561), and atosiban groups (-0.30 ± 3.66 pg/ml, *p* < 0.001, *d* = 1.202). The OT changes in the PLC group were significantly lower than in the 48IU OT *(p* < 0.001, *d* =1.139), 24IU OT (*p* = 0.001, *d* =0.595), and atosiban + 24IU OT groups (*p* = 0.005, *d* =0.579) (see Fig 3a). The post-treatment changes in OT concentrations were also significantly positively correlated with time spent viewing the social stimuli in both OT treatment groups (Pearson: 24IU OT *r* = 0.652, *p* < 0.001, 95% *CI* = [0.471,0.842]; 48IU OT *r* = 0.437, *p* = 0.003, 95% *CI* = [0.197,0.603]) and in the PLC group (*r* = 0.341, *p* = 0.022, 95% *CI* = [0.106,0.543]), but not in the atosiban (*r* = 0.044, *p* = 0.784, 95% *CI* = [-0.251,0.361]) or atosiban + 24IU OT (*r* = -0.080, *p* = 0.625, 95% *CI* = [-0.419,0.229]) groups. Fig 3b shows the scatter plots and linear regression fit lines for the 48IU OT and 24IU OT groups. The corresponding plot for the PLC group is provided in Supplementary Fig S1. If we combined the two OT-treatment groups there was also a robust overall positive correlation with time spent viewing the social stimuli (*r* = 0.59, *p* < 0.001). There was also a smaller significant correlation if we only considered absolute OT concentrations at T3 without considering the change from baseline (*r* = 0.32, *p* = 0.001). There was no correlation between basal OT concentrations and the magnitude of increases in T3 in the 48IU OT group (*r* = -0.041, *p* = 0.798, 95% *CI* = [-0.398,0.230]). Thus, in the absence of atosiban treatment, there was a consistent positive correlation between the magnitude of post-treatment increases in plasma OT concentrations and the amount of time spent viewing social stimuli. Given the paradigm design where either only social or geometric stimuli were viewed there was a corresponding pattern of opposite correlations between changes in OT concentrations and geometric stimuli (see supplementary results).

**Fig 3.**
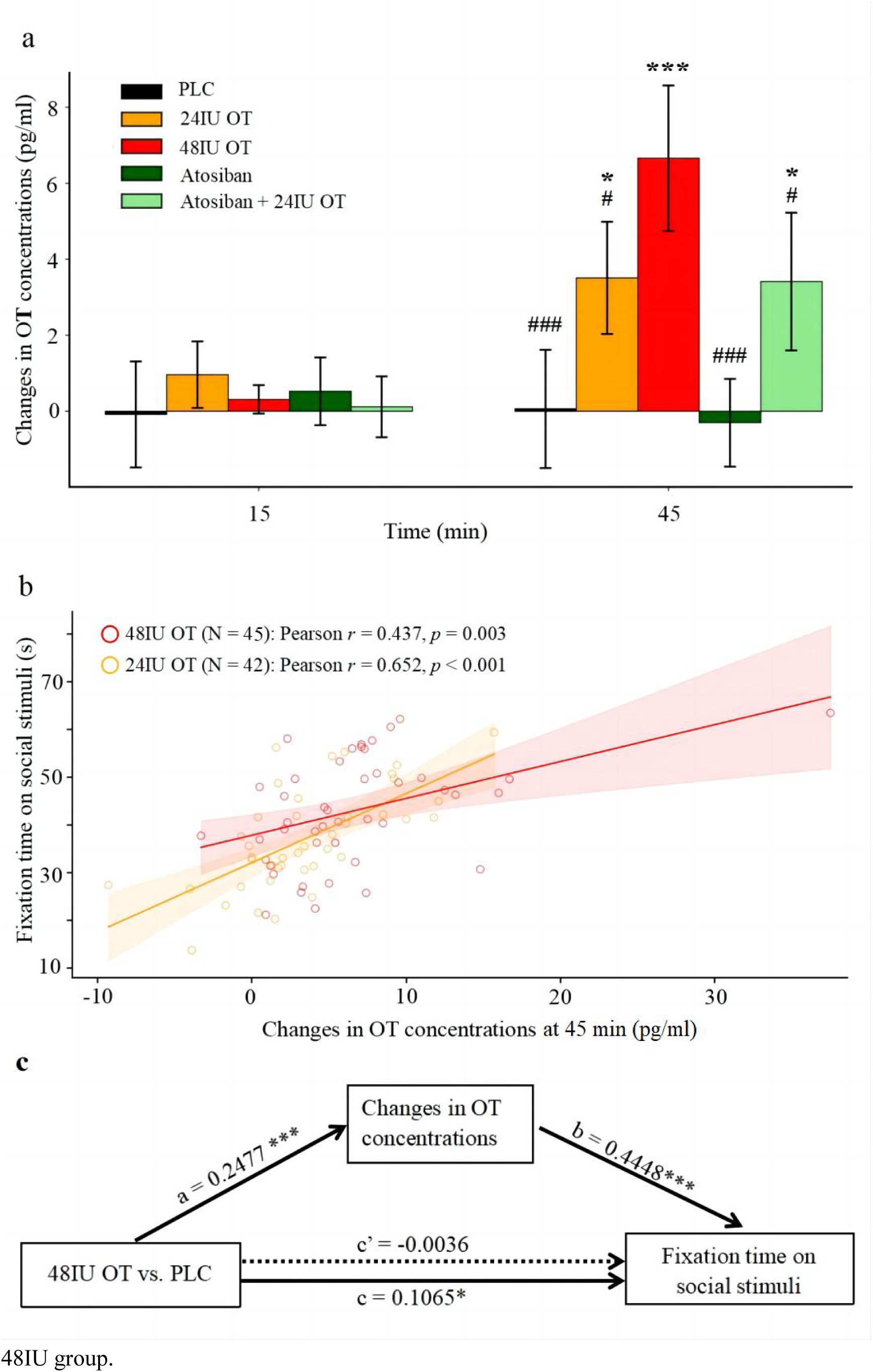
Treatment effects on plasma OT concentrations and associations with preference for social stimuli. **a)** Altered plasma OT concentrations across OT and atosiban treatment groups at +15 (T2) and +45 (T3) min time points compared to time 0 (T1). \**p* < 0.05, ** *p* < 0.01, *** *p* < 0.001 compared to the PLC group. #*p* < 0.05, ## *p* < 0.01, ### *p* < 0.001 compared to the 48IU OT group. Mean ± SD OT concentrations at T1 were 10.57 ± 5.08 pg/ml. **b)** Positive correlations (Pearson) between altered plasma OT concentrations at the 45 min time point (T3) relative to T1 time points and fixation time on social stimuli in 24IU OT and 48IU OT groups. **c)** Mediation analyses showing that increased changes in OT concentrations significantly mediated differences between the PLC and 48IU OT treatment groups on fixation time on social stimuli. \**p* < 0.05, *** *p* < 0.001.

### Mediation analysis of the influence of OT changes on preference for social stimuli

A mediation analysis showed that the OT concentration changes significantly mediated the difference between the PLC and 48IU OT groups in fixation time on social stimuli (indirect effect = 0.1102, *SE* = 0.0259, 95% *CI* = [0.0627,0.1658], bootstrap = 5,000, *p* < 0.001, see Fig. 3c).

### Correlations between trait autism scores and social attention across treatment groups

As hypothesized, there was a negative correlation between total AQ scores and time spent viewing the social (dancing individuals) stimuli only in the 48IU OT group (Pearson *r* = -0.256, *p* = 0.043 one-tailed, 95% *CI* = [-1.00, -0.011], see Fig 4) but not in the other groups (*rs* = -0.104 - 0.103, *ps* = 0.246 - 0.438 one-tailed).

**Fig 4.**
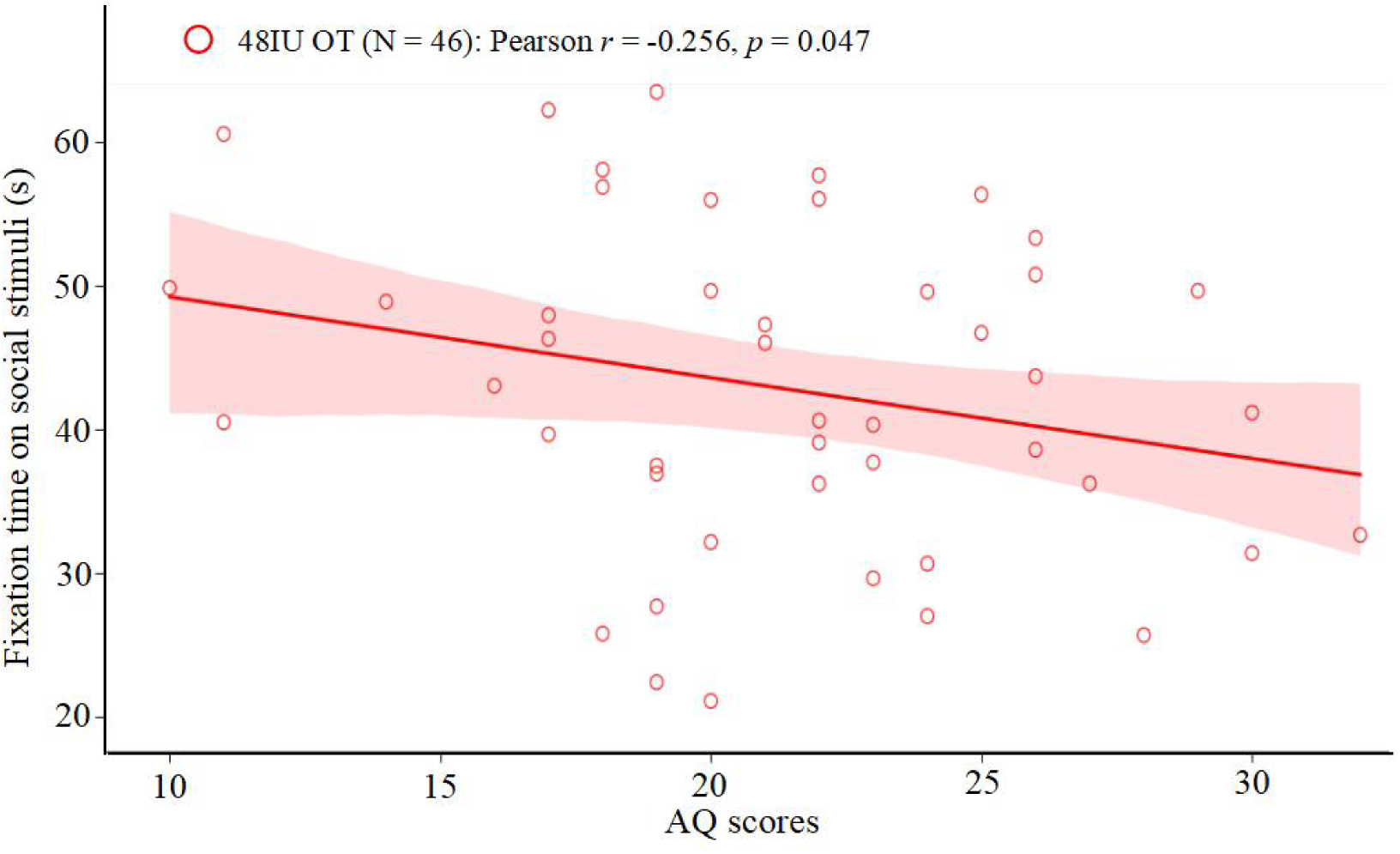
Associations between trait autism scores and preference for social stimuli. Negative correlation (Pearson, one-tailed) between AQ scores and fixation time on social stimuli in 48IU OT group.

## Discussion

The current study used eye-tracking in combination with a pharmacological approach to establish the importance of basal and increased peripheral OT concentrations acting on peripheral receptors on attention to social as opposed to non-social stimuli. A dynamic social versus geometric pattern presentation paradigm was employed which is increasingly used in the context of diagnosis and therapeutics in autism (Kou et al., 2019; Moore et al., 2018; Pierce et al., 2016; Le et al., 2020; Le et al., 2022). Results showed that a 48IU, but not 24IU, dose of OT significantly increased gaze towards social stimuli and correspondingly reduced it towards non-social geometric stimuli. Gaze durations towards social stimuli in the 48IU OT group were also negatively associated with AQ score. The OT receptor antagonist atosiban had the opposite effect to OT, and when given prior to 24IU OT still reduced gaze towards social stimuli and tended to increase it towards non-social geometric patterns. OT dose-dependently increased plasma OT concentrations 30 minutes post-administration, whereas, in line with a previous study (Babic et al., 2015), atosiban had no effect, and the magnitude of OT increases was positively correlated with the time spent viewing social stimuli. Overall, findings therefore provide support for both endogenous and exogenous peripheral OT acting to facilitate attention towards social cues, possibly via its receptors in cardiac and gastrointestinal systems influencing vagal projections to the brain (see Yao and Kendrick, 2025) and in support of Porges polyvagal theory (Porges, 2021).

Oromucosal OT dose-dependently increased gaze duration towards the dynamic social stimuli and correspondingly reduced it towards the dynamic, non-social geometric pattern stimuli. Further analysis of the number of fixation counts indicated that changes in these were mainly contributing to the effects of OT on overall fixation duration. Since oromucosal administration of OT is unlikely to directly enter the brain, this finding contributes to growing evidence that exogenous administration of OT may primarily produce functional effects by acting on its peripheral receptors (Yao & Kendrick, 2025). Only the 48IU dose produced significant effects and it also increased plasma OT concentrations to a greater extent than the 24IU dose 30 min after administration. While the half-life of OT in blood is likely to be less than 6 minutes (Uvnäs-Moberg 2024) with high doses of oromucosal administration absorption is slow and prolonged with a Tmax of 34 minutes (Xu D et al., 2024) and concentrations remain increased for a further 30-45 minutes (Kou et al., 2021; Xu D et al., 2024). Furthermore, significantly increased concentrations in the mouth (saliva) can still be seen 2h after administration (Xu D et al., 2024). However, it is also possible that concentrations are further boosted by increased endogenous release as a result of positive feedback following vagal stimulation (Zhu et al., 2022). We had expected the 24IU dose would also produce a significant effect on increasing preference for the social stimuli in line with our previous study using a 24IU intranasal dose (Le et al., 2020). However, oromucosal OT administration is significantly less effective at increasing peripheral OT concentrations than intranasal administration (Xu et al., 2022; Xu D et al., 2024). Indeed, in the current study plasma OT concentrations were only increased by 3.51 pg/ml with 24IU OT, and 3.41pg/ml in the atosiban + 24IU OT group, and 6.66 pg/ml in the 48IU OT group, compared to 9.7 pg/ml 30 min after intranasal administration of 24IU OT (Xu et al., 2022). The close relationship between increased plasma OT concentrations and time spent viewing the social stimuli was evidenced by positive correlations not only in the 48IU OT group but also in the 24IU OT and the two OT-treatment groups combined. There was even a significant correlation in the PLC group despite only small mean changes in endogenous OT, but not in the two groups treated with atosiban. The lack of an overall significant treatment effect in the 24IU group despite the correlation probably reflects greater variation across subjects with some showing effects but others none at all, whereas in the 48IU group subjects showed more consistent changes which were also correlated with changes in OT. The greater time spent viewing the social stimuli in the 48IU OT vs PLC group was also mediated by the increase in plasma OT concentrations. Overall, these findings suggest an association between increased (either exogenous or endogenous) peripheral OT concentrations and social attention in individuals and that at higher doses of oromucosal OT administration can significantly facilitate social attention across individuals and may therefore have potential therapeutic use in disorders with social interaction and communication problems such as autism. As hypothesized, oromucosally administered atosiban had the opposite effect to OT by reducing the time spent viewing dynamic social stimuli, although a corresponding increase in time viewing the non-social dynamic geometric patterns only achieved significance compared with 48IU OT. Thus, preventing peripheral OT receptors from responding to endogenous OT may reduce visual attention to social stimuli and indicates that reduced basal concentrations of the peptide in psychiatric disorders (Moerkerke et al., 2021; Xie et al., 2021; Ferreira et al., 2022) may contribute towards reduced interest in social stimuli. Atosiban administration in humans may therefore provide a useful tool to establish the importance of endogenous concentrations of OT. Given atosiban only acts as an antagonist of OT receptor G(q)-coupling, and is a partial agonist on G(i)-coupling (Reversi et al., 2005; Ślusarz et al., 2004), the facilitatory effects of OT on social attention are likely to be mediated via G(q)-coupled receptors. Indeed, a recent study in rodents found that OT and atosiban produced similar rather than opposite anxiolytic effects following chronic social defeat stress suggesting that both were acting via G(i)-coupled receptors in this context (Canto-de-Souza et al., 2025).

While we cannot completely rule out the possibility that atosiban given orally might cross the blood-brain-barrier to influence brain OT receptors, this is unlikely given that it is relatively impermeable to it, similar to OT (Marshall et al., 2007; Babic et al., 2015). As discussed above, atosiban may be exerting its effects via preventing OT from binding to its peripheral G(q)-coupled receptors in the cardiovascular and gastrointestinal systems which may contribute to vagal modulation of brain function (Yao & Kendrick, 2025; Nowacka et al., 2025) in support of polyvagal theory (Porges et al., 2019). Indeed, the peripheral receptors in these systems primarily signal through G[q]-coupled receptors (Babic et al., 2015), although possibly at higher doses of atosiban might have produced different effects, potentially via agonist effects on G(i)-coupled receptors. There was at least some evidence that the chosen dose of 150μg was sufficient to overcome any effects of 24IU OT, since the pattern of gaze durations for social stimuli was significantly different from when 24IU OT was given alone and not different from that observed after atosiban alone. Atosiban also prevented the correlation between increased OT concentrations and time spent viewing social stimuli seen with 24IU OT alone.

As predicted, we found a significant negative association between trait autism (AQ scores) and time spent viewing the social stimuli in the 48IU OT group, but none of the other treatment groups. This suggests that, acute exogenous OT treatment may produce greater effects in individuals with the lower rather than higher AQ scores. This is in agreement with the findings of our previous within-subject design study using intranasal OT (Le et al., 2020). However, with repeated dosing, OT has been shown to increase the amount of time autistic children view the social stimuli in this paradigm (Le et al., 2022) and therefore it may require multiple doses of OT to have robust effects on individuals with greater autistic symptoms.

While some studies have reported anxiolytic effects of OT, particularly in animal models, this is less established in humans (Yao and Kendrick, 2025) and we found no evidence for reductions in anxiety in line with the previous study using intranasal OT in this same (Le et al., 2022) and other paradigms (Rashidi et al., 2025). These findings make it unlikely that non-specific effects of OT on stress contributed to our findings.

The current study has several limitations. Firstly, only male subjects were included due to potential concerns about using atosiban in females. There is evidence that OT can have sex-dependent effects (Yao & Kendrick, 2025), so findings would need to be confirmed in females. Secondly, different effects might have occurred with higher doses of atosiban, although it did produce significant effects alone, and in combination with 24IU OT it significantly reduced time spent viewing social stimuli relative to 24IU OT alone. Clearly it will be important however to assess its antagonistic effects when given prior to a higher dose of OT which significantly increases viewing of social stimuli. Thirdly, we cannot rule out the possibility that OT was acting via V1a receptors rather than OT receptors given that there can be cross talk between them (Song & Albers, 2018). However, OT has at least a 10 to100-fold lower affinity for V1a receptors and while OT and vasopressin can have similar effects on aspects of social behavior they can also exert different functional effects on brain and behavior (Zhao et al., 2026). Finally, although we hypothesize that OT and atosiban are acting primarily via peripheral receptors and vagal projections to the brain, we cannot completely rule out the possibility that some oromucosal OT might have entered the brain directly via the trigeminal nerve. However, this would involve both substances travelling a great distance from the tongue to the brain and probably take too long to account for functional effects observed starting at 30 minutes after administration (Johnson et al., 2010). OT, although not atosiban, might also bind to RAGE, and cross the blood brain barrier although this has not yet been demonstrated in humans.

Overall, findings of the current study provide evidence for opposite effects of OT and atosiban on preference for viewing dynamic social as opposed to non-social geometric patterns. This underlines the importance of both basal endogenous concentrations of OT and increased ones following exogenous OT administration for facilitating social attention. Given that both OT and atosiban administered via an oromucosal route are likely to primarily influence peripheral OT receptors, the findings further suggest that many functional effects of OT are mediated via its actions on peripheral receptors subsequently influencing brain function, possibly via G(q)-coupled receptors in the vagal system.

## Acknowledgements

The research was supported by Sichuan Province Key Research and Development Project [grant number 2023YFWZ0003 and Key Technological Projects of Guangdong Province “Development of New Tools for Diagnosis and Treatment of Autism” grant number 2018B030335001

## Author contributions

KMK. and YZ designed the study; YZ conducted the experiment and collected the data; HL completed the ELISA analysis; XW completed the drug distribution during the double blinded process; YW provided technical assistance and supervision; YZ and KMK performed the data analysis; YZ wrote the manuscript draft; KMK critically revised the manuscript and all authors approved the final draft.

## Data availability

The data that support the findings of this study are available from the corresponding author upon reasonable request.

## Abbreviations

ANOVA: analysis of variance
AOI: areas of interest
AQ: Autism Spectrum Quotient
BDI-Ⅱ: Beck depression inventory-Ⅱ
DSG: dynamic social versus geometric stimuli paradigm
ELISA: enzyme-linked immunosorbent assay
HSD: honestly significant difference
IU: International Unit
LSAS: Leibowitz social anxiety scale
OT: Oxytocin
PLC: placebo
RAGE: advanced glycation end-products
STAI: state-trait anxiety inventory
Tmax: time to maximum plasma concentration
V1aR: V1a receptor
V2R: V2 receptor.

