## Supplementary for "Oromucosal administration of oxytocin and its receptor antagonist atosiban respectively increase and decrease preferential attention to dynamic social stimuli in men"

**Supplementary methods**

*Test of effective blinding of treatment groups*

The majority of subjects (89.6%) did not report consistent taste differences between sprays and all treatments included glycerine which gives a slightly sweet taste. We did record participants' subjective sensory feedback regarding the study medications after the experimental procedure too. Since our recruitment advertisement stated that participants would receive either one or two of the three medications (placebo / OT / atosiban), we asked a standardized question after they completed the experiment: "What do you think you received for the first and the second spray?"

We conducted stratified analyses based on five independent group designs, rather than simply pooling all participants together. Since each participant made a three-choice guess (placebo / OT / atosiban) for each of the two sprays after the experiment, the theoretical probability of guessing both correctly by chance is: P = 1/3 × 1/3 = 1/9 ≈ 11.11%. To evaluate the blinding integrity, we performed statistical analyses at two levels.

First, within-group analysis. Using the theoretical chance probability of 1/9 as a reference, we conducted one-sample binomial tests (one-tailed, testing whether the observed proportion < 1/9) separately for each of the five groups to examine whether the observed proportion of participants who correctly identified both sprays deviated from this theoretical value. The correct rates in PLC + PLC (5 correct, one-sample binomial test, one-tailed, *p* = 0.515), PLC + 24IU OT (2 correct, *p* = 0.073), and atosiban +24IU OT (2 correct, *p* = 0.073) did not differ significantly from the chance level, while the correct rates in atosiban + PLC and PLC + 48IU OT groups were significantly lower than chance (only 1 participant in each group guessed both drugs correctly, *ps* = 0.020). The above results indicated that participants in all treatment groups were unable to identify their assigned treatments through guessing.

Second, between-group analysis. We performed Fisher's exact test on a 5 × 2 contingency table (group × guess correct/incorrect) to examine whether the correct rates differed significantly across the five groups. The results showed no significant difference in correct rates among the five groups ( χ^2^ = 0.4082, *p* = 0.432), indicating that participants' guessing accuracy was independent of their actual group assignment.

**Supplementary results**

*Comparisons of fixation counts across the different treatment groups*

**Table S1. Comparisons of fixation counts between different treatment groups**

| Group | N | Mean fixation counts (%) | |
| --- | --- | --- | --- |
|  |  | Social stimuli | Geometric stimuli |
| PLC | 46 | 61.59 ± 12.69 **^†^** | 35.09 ± 12.49 ^#^ |
| 24IU OT | 45 | 60.23 ± 14.34 ^#^ | 35.24 ± 13.14 ^#^ |
| 48IU OT | 46 | 66.71 ± 14.09 | 29.22 ± 14.19 * |
| Atosiban | 45 | 55.23 ± 15.64 *^###^ | 39.67 ± 16.47 ^##^ |
| Atosiban + 24IU OT | 45 | 54.92 ± 13.76 *^###^ | 40.15 ± 13.97^###^ |

Note: *p < 0.05, ** p <0.01, *** p < 0.001 compared to the same stimuli in the PLC group;† p = 0.068, # p < 0.05, ## p < 0.01, ### p < 0.001 compared to the same stimuli in the 48IU group.

*Correlations between changes in OT concentration and time spent viewing the geometric stimuli*

As expected, given the design of the paradigm where subjects could only view two different stimuli, the pattern of correlations between the change in plasma OT and time spent viewing the non-social geometric stimuli was the opposite and of similar magnitude to that for time spent viewing the social stimuli. Pearsson correlation was significant in both the 24 IU OT group (r = -0.620, p < 0.001) and the 48 IU OT group (r = -0.378, p = 0.011), although not in the placebo group (r = -0.228, p = 0.131) or the atosiban (r = 0.030, p = 0.853) and atosiban + 24IU OT groups (r = 0.135, p = 0.412).

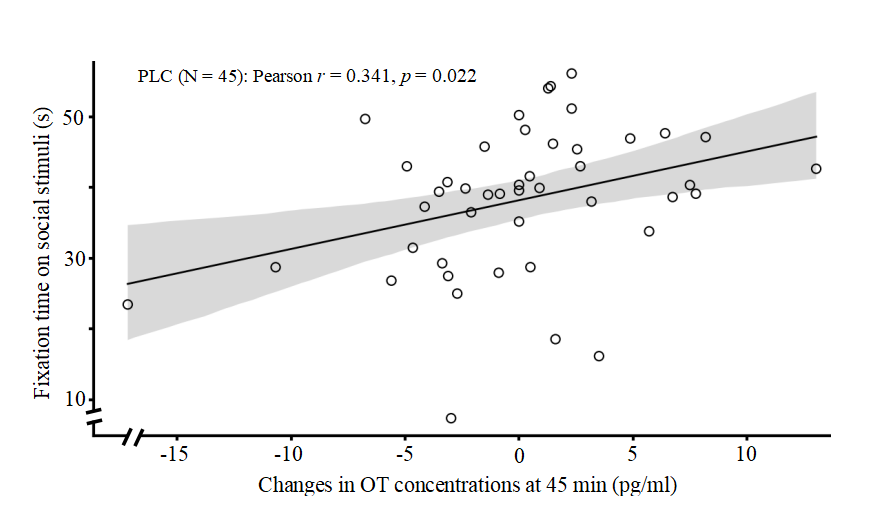

**Figure S1.** Scatter plot showing the association between time spent viewing the social stimuli and absolute change in plasma OT concentrations from time 0 (T1) to 45 (T3) minutes later
